# No immunological interference or safety concerns when adjuvanted recombinant zoster vaccine is coadministered with a COVID-19 mRNA-1273 booster vaccine in adults aged 50 years and older: A randomized trial

**DOI:** 10.1101/2023.03.10.23286967

**Authors:** Abdi Naficy, Adrienne Kuxhausen, Paola Pirrotta, Brett Leav, Jacqueline Miller, Kate Anteyi, Mark S Adams, Jasur Danier, Thomas Breuer, Agnes Mwakingwe-Omari

## Abstract

**Background:** There is growing consensus that COVID-19 booster vaccines may be coadministered with other age-appropriate vaccines. Adding to the limited available data supporting coadministration, especially with adjuvanted vaccines, could enhance vaccine coverage in adults.

**Methods:** In this phase 3, randomized, open-label study, eligible adults aged ≥50 years were randomly assigned (1:1) to receive mRNA-1273 (50µg) booster vaccination and a first dose of recombinant zoster vaccine (RZV1) 2 weeks apart (Seq group) or concomitantly (Coad group). The second RZV dose (RZV2) was administered 2 months post- RZV1 in both groups. Primary objectives were noninferiority of anti-glycoprotein E and anti-Spike protein antibody responses in the Coad group compared to the Seq group. Safety and further immunogenicity assessments were secondary objectives.

**Results:** 273 participants were randomized to the Seq group, 272 to the Coad group. Protocol-specified non-inferiority criteria were met. The adjusted geometric mean concentration ratio (Seq/Coad) was 1.01 (95% confidence interval [CI], 0.89–1.13) for anti- gE antibodies 1-month post-RZV2, and 1.09 (95% CI 0.90–1.32) for anti-Spike antibodies 1- month post-mRNA-1273 booster. No clinically relevant differences were observed in overall frequency, intensity, or duration of adverse events between the 2 study groups. Most solicited adverse events were mild/moderate in intensity, each with median duration ≤2.5 days.

Administration site pain and myalgia were the most frequently reported in both groups.

**Conclusions:** Coadministration of mRNA-1273 booster vaccine with RZV in adults aged ≥50 years was immunologically noninferior to sequential administration and had a safety and reactogenicity profile consistent with both vaccines administered sequentially (clinicaltrials.gov NCT05047770).

**Visual abstract:** 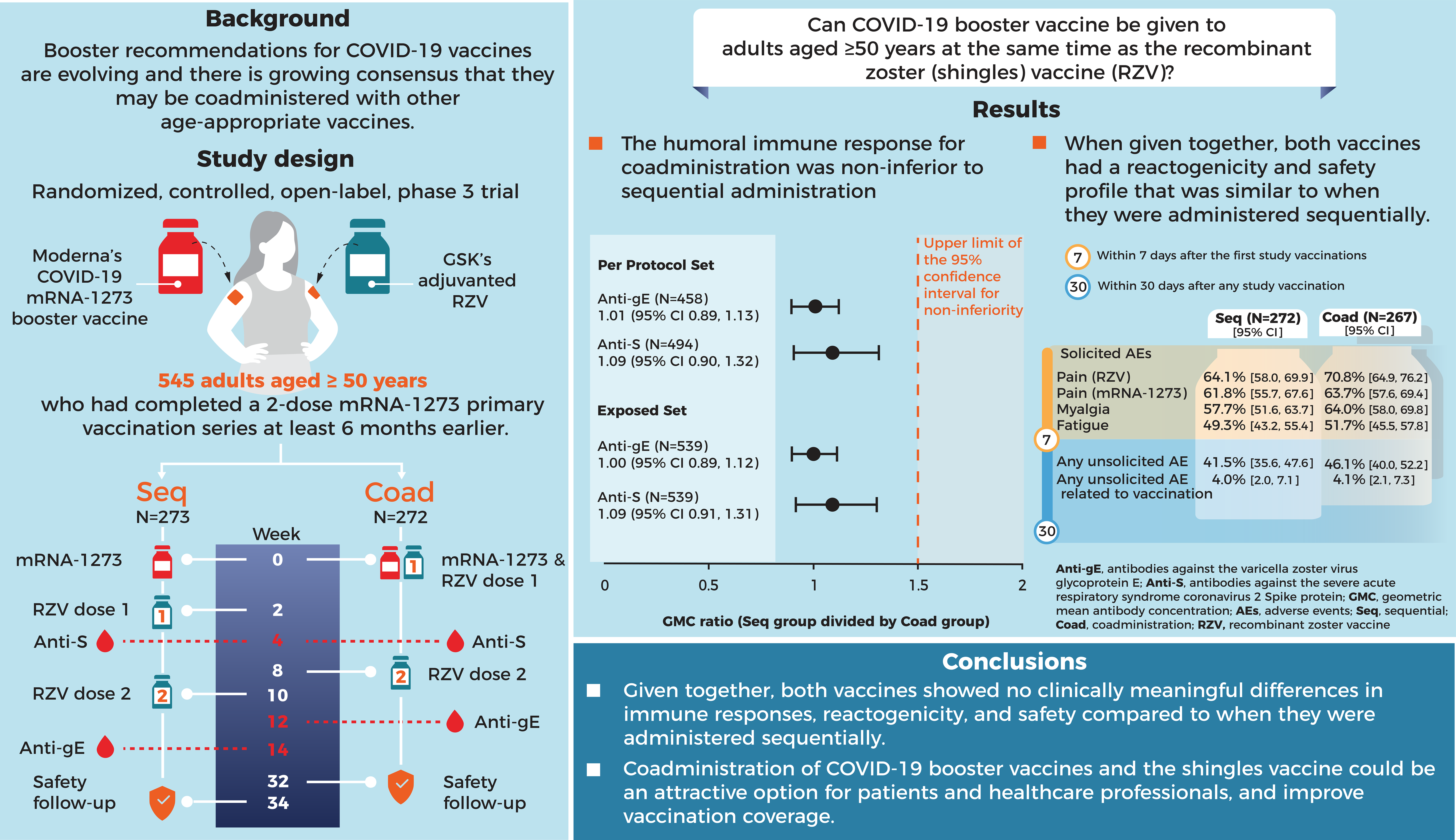

## INTRODUCTION

Accumulating real-world data substantiate the protective benefits of mRNA coronavirus disease 2019 (COVID-19) vaccines and the need for additional doses beyond the primary series due to waning immunity and/or emergence of new variants.^1, 2^ Booster recommendations for COVID-19 vaccines are evolving and there is growing consensus that they may be coadministered with other age-appropriate vaccines.^3–5^ During the COVID-19 pandemic, vaccination of adults reached historic highs, with 69% of the global population having received at least 1 dose of COVID-19 vaccine as of December 2022.^6^ However, rates for other vaccines were negatively impacted.^7^ Maintaining and improving uptake of COVID- 19 booster vaccines remains an important public health priority in many countries. It is also important to continue to improve the uptake of other vaccines routinely recommended for adults. In this context, vaccine uptake and coverage would be positively impacted if vaccines recommended for adults could be coadministered with COVID-19 booster vaccines.

Numerous health agencies including the United States of America (US) Centers for Disease Control and Prevention (CDC) have recommended, in the absence of specific contraindications, administration of COVID-19 booster vaccines on the same day as other vaccines.^8–11^ To date, clinical trial data describing safety and immunogenicity of coadministration with COVID-19 vaccines are limited to seasonal influenza vaccines.^12, 13^

In the context of coadministration of COVID-19 vaccines with other vaccines, the CDC recommends to consider the reactogenicity profile of the vaccines and that it is unknown whether reactogenicity of COVID-19 vaccine is increased with coadministration, including with other vaccines known to be more reactogenic, such as adjuvanted vaccines.^14^ In addition, the United Kingdom guidance currently states that because of the potential difficulty of attributing systemic side-effects to the adjuvanted shingles vaccine, a 7-day interval should ideally be observed.^9^ Overcoming barriers to coadministration will require the provision of information to health agencies, healthcare providers, and the general public about the benefits versus risks of coadministration, supported by clinical trial data.

The adjuvanted recombinant zoster vaccine (RZV; *Shingrix*, GSK) is a non-live subunit vaccine that contains the varicella-zoster-virus glycoprotein E (gE) as the active ingredient, together with the liposome-based adjuvant system AS01_B_. In clinical trials, RZV demonstrated high efficacy in preventing herpes zoster (HZ), 97.2% and 91.3% in adults aged ≥50 years and ≥70 years, respectively.^15, 16^ RZV also demonstrated unprecedented efficacy against HZ of 68.2% in autologous hematopoietic stem cell transplant patients.^17^ RZV has been approved in over 40 countries worldwide, including the US and European Economic Area, for the prevention of HZ in adults aged ≥50 years and in adults aged ≥18 years who are at increased risk of HZ.^18, 19^ These populations are similar to those at high risk of severe COVID-19 and its complications.^20^

Moderna’s COVID-19 vaccine, mRNA (mRNA-1273) is an mRNA-based vaccine encapsulated in a lipid nanoparticle. The vaccine includes a single mRNA sequence encoding the pre-fusion stabilized Spike (S) protein of severe acute respiratory syndrome coronavirus 2 (SARS-CoV-2), Wuhan strain. Two doses of 100 µg mRNA-1273, as a primary series, showed 94.1% efficacy at preventing COVID-19, including severe disease,^21^ and was approved as *Spikevax* (Moderna) for the prevention of COVID-19 in individuals aged ≥18 years in the US and aged ≥6 months in the European Union (EU).^21, 22^ A booster dose (50 ug) of mRNA-1273 for the prevention of COVID-19 received an initial Emergency Use Authorization in the US in December 2021 for adults aged ≥18 years, and was approved in the EU for individuals aged ≥12 years.^22, 23^

Although coadministration with RZV is not contraindicated, no clinical trial data is available on its coadministration with COVID-19 vaccines. To address this data gap and provide evidence-based guidance for healthcare providers making decisions on such vaccine coadministrations, we conducted a clinical trial to assess the safety and immunogenicity of coadministration of a booster dose (50 µg) of mRNA-1273 with either seasonal quadrivalent influenza vaccine in adults aged ≥18 years, or RZV in adults aged ≥50 years. The results of coadministration of RZV and mRNA-1273 are reported here.

## METHODS

### Study design and participants

This was a phase 3, randomized, open-label, multi-center clinical trial conducted in the US. Eligible adults were randomly assigned (1:1) to receive coadministration of a booster dose (50 µg) of mRNA-1273 and the first dose of RZV (RZV1) (Coad group), or the mRNA-1273 booster followed 2 weeks later by RZV1 (Sequential [Seq] group). All study participants received the second dose of RZV (RZV2) 2-months post-RZV1 and were followed for safety endpoints until 6-months post-RZV2 (clinicaltrials.gov NCT05047770).

Eligible adults aged ≥50 years were healthy or medically stable who had completed a 2-dose mRNA-1273 primary vaccination series at least 6 months prior to study vaccination. A full list of eligibility criteria is provided in the Supplement.

The study was conducted according to Good Clinical Practice guidelines and the protocol was approved by all applicable institutional review boards (Advarra Institutional Review Board, Western Copernicus Group [WCG] Institutional Review Board). Written informed consent was obtained from each participant prior to enrolment.

### Randomization

Participants were stratified by age (50–59, 60–69, ≥70 years) and centrally randomized to either the Seq or Coad group. The randomization system allocated a participant identification number and provided the treatment number to be administered.

### Objectives

The primary objectives were (1) to demonstrate noninferiority in terms of humoral immunogenicity of 2 doses of RZV when RZV1 was coadministered with an mRNA-1273 booster dose compared to RZV1 administered 2 weeks after mRNA-1273; and (2) to demonstrate noninferiority in terms of humoral immunogenicity of a booster dose of mRNA- 1273 when coadministered with RZV1 compared to its administration 2 weeks prior to RZV1. Secondary objectives were to characterize the immune responses to RZV and mRNA- 1273, and to evaluate safety and reactogenicity of the study vaccines, and are provided in the Supplement.

### Study interventions and procedures

The composition of RZV and mRNA-1273 is provided in the Supplement. Blood samples (15 ml) were collected prior to each vaccination, 4 weeks post-mRNA-1273 administration, and 4 weeks post-RZV2.

Anti-gE antibodies were measured using an enzyme-linked immunosorbent assay at GSK.^24^ SARS-CoV-2 anti-S IgG antibodies were measured using a Multiplex Electrochemiluminescence assay at PPD Laboratory Services.

Solicited local and systemic adverse events (AEs) with onset within 7 days after each vaccination were recorded using electronic diaries. Unsolicited AEs were recorded for 30 days after each vaccination. Serious adverse events (SAEs), intercurrent medical conditions, potential immune-mediated diseases (pIMDs), pregnancies, AEs of special interest (AESIs), and cases of COVID-19 and HZ were collected up to 6 months after RZV2. All solicited AEs were considered causally related to study vaccination. Causal relationship to vaccination of all other AEs was assessed by the investigators and independently by the sponsor. A joint safety review team with GSK and Moderna representatives oversaw participant safety.

Randomization was temporarily paused as per protocol after 10% of participants were vaccinated and safety data collected for 7-days post-vaccination.

The protocol-defined list of AESIs, AE intensity grading table, causality assessment criteria by the investigator, and protocol-defined study holding rules are provided in the Supplement.

### Statistical analysis

The Exposed Set included all participants who received at least 1 dose of a study vaccine. The Per-Protocol Set (PPS) included study participants who met eligibility criteria, received all vaccinations according to their random assignment, complied with protocol-defined procedures, did not receive prohibited medications or vaccines, had available post- vaccination immunogenicity data, and where the administration site was known.

Anti-gE antibody concentrations and anti-S protein antibody concentrations were expressed as between-group ratios of the geometric mean concentration (GMC) 1 month post-RZV2 and 1 month post-mRNA-1273 booster, respectively. The 95% CIs of the between-group GMC ratios were computed using an analysis of covariance model on the log_10_ transformation of the concentrations. The age strata and prevaccination log-transformed antibody concentrations were covariates, and vaccine group was a fixed effect.

GMCs were calculated by taking the anti-log of the mean of the log concentration transformations. Noninferiority of the anti-gE antibody or anti-S antibody response was demonstrated if the upper limit of the 95% CI of the adjusted GMC ratio (Seq over Coad) was <1.5, 1-month post-RZV2 or post-mRNA-1273 booster, respectively.

Assuming a GMC ratio of 1.1 between the Seq and Coad groups, the global power to meet both co-primary objectives with 245 evaluable participants in the Seq and Coad groups was 90%. Assuming that about 10% of the randomized participants would not be evaluable, approximately 546 participants (273 in each study group) were planned.

Descriptive immunogenicity analyses were also performed. Safety was evaluated descriptively, without pre-defined statistical criteria. All statistical analyses were performed using SAS version 9.4.

## RESULTS

### Participants

The study was conducted between 7 October 2021 and 29 August 2022 at 47 sites in the US. From a total of 545 participants randomized, 539 were vaccinated (Exposed Set; 272 in the Seq and 267 in the Coad group), and 91.2% in the Seq and 92.6% in the Coad group completed the study (**Fig. 1**).

**Figure 1.**
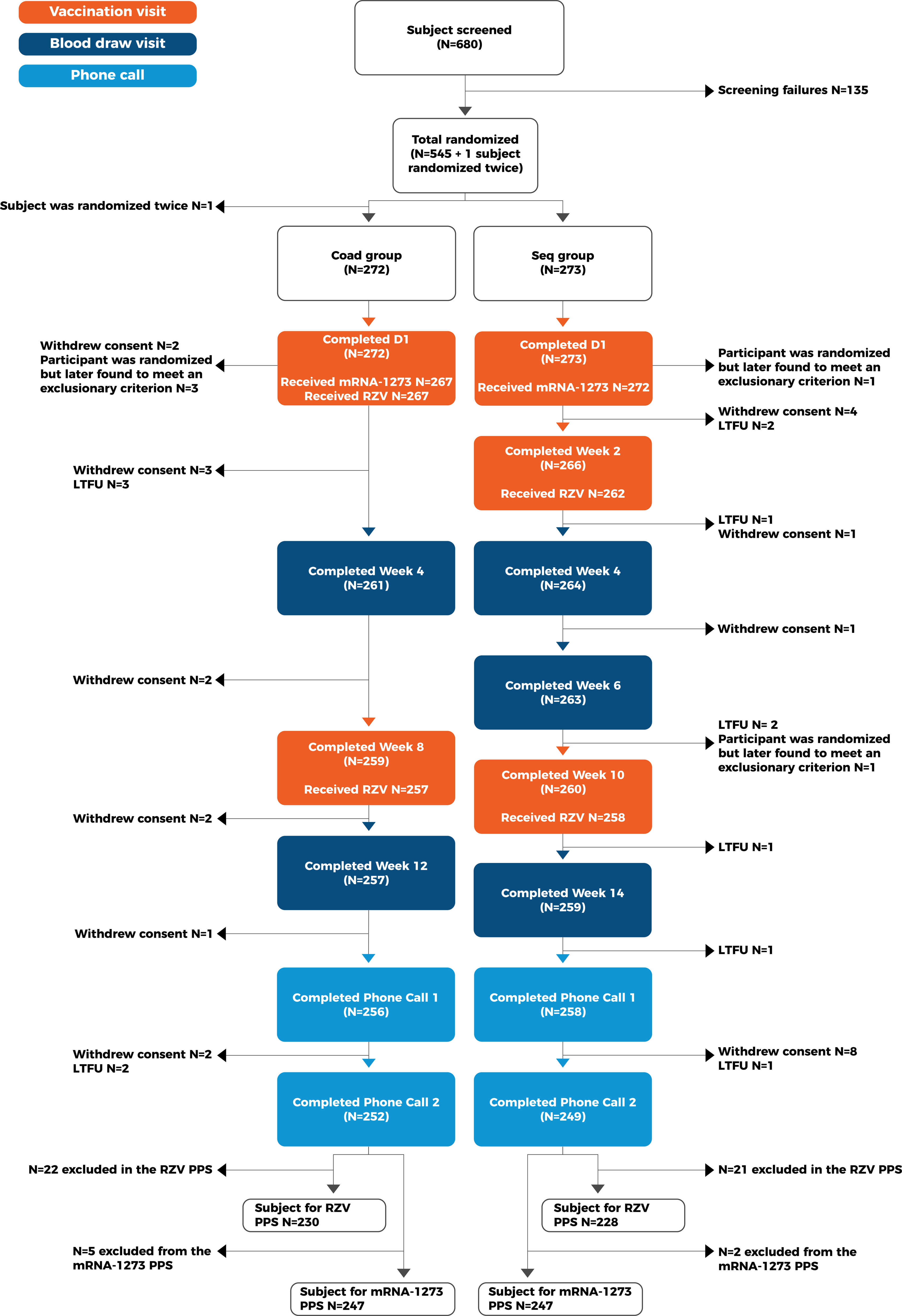
Participant flow D, study day; LTFU, lost to follow-up; mRNA-1273, Moderna’s mRNA COVID-19 vaccine; N, number of participants, PPS, per protocol set; RZV, recombinant zoster vaccine Seq group received the mRNA-1273 booster dose followed 2 weeks later by the first dose of RZV. Coad group received coadministration of the mRNA-1273 booster and the first dose of RZV

The study groups were well balanced in terms of demography (**Table 1**). The median age of participants was 61 years (range 50–88 years) and 22% were aged ≥70 years, 56% were female, 92% were white, 4.6% were black, and 22% were Hispanic.

**Table 1.**
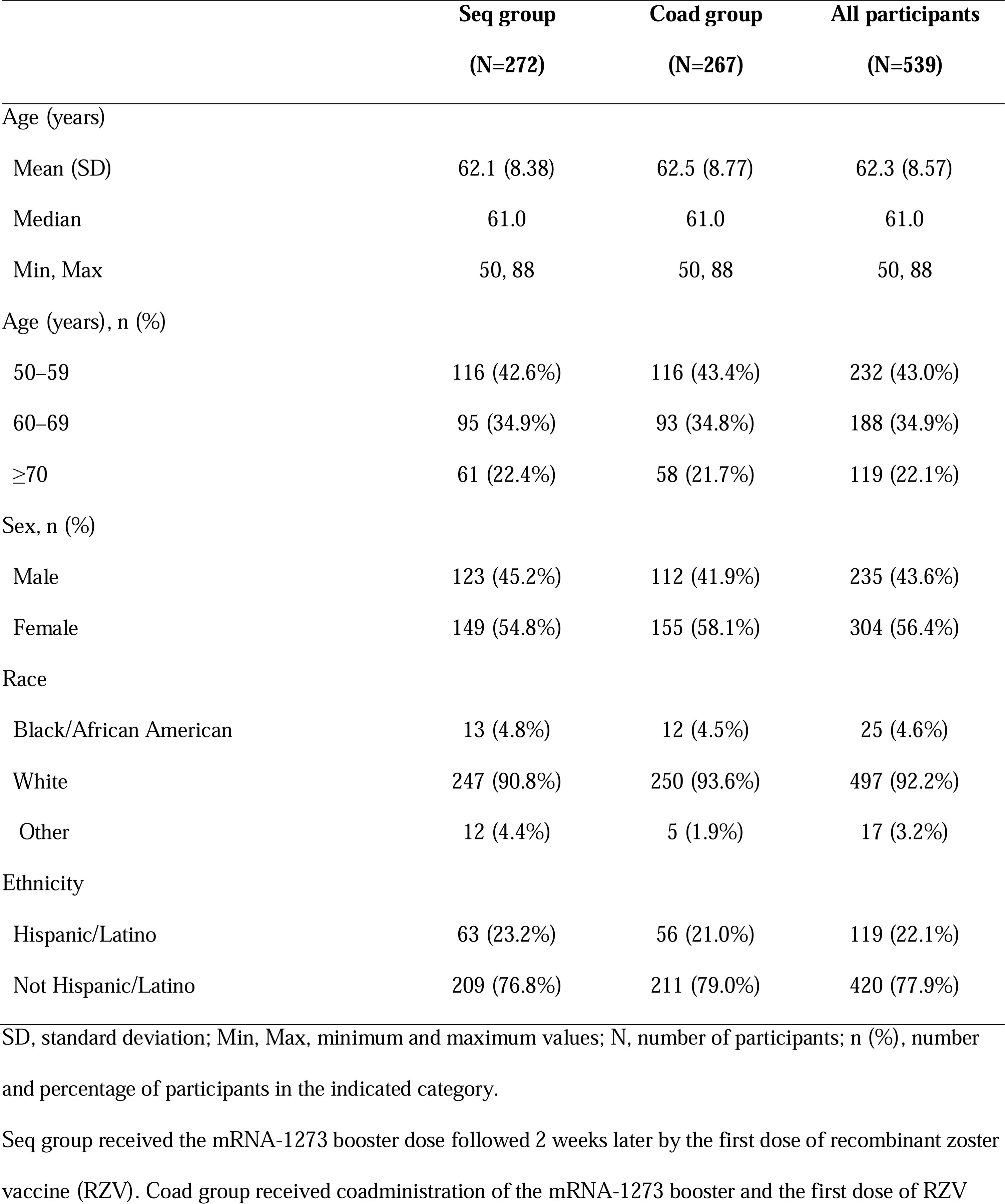
Demographic characteristics of study participants (Exposed Set)

### Immunogenicity results

Noninferiority of the humoral immune response to the gE and S antigens was demonstrated according to the protocol-specified criteria. For the PPS, the adjusted GMC ratio (Seq over Coad) was 1.01 (95% CI, 0.89–1.13) for anti-gE antibodies 1-month post-RZV2 (**Table 2**), and 1.09 (95% CI, 0.90–1.32) for anti-S antibodies 1-month post-mRNA-1273 booster (**Table 3**). In a secondary analysis on the Exposed Set, the corresponding GMC ratios were 1.00 (95% CI, 0.89–1.12) for anti-gE antibodies and 1.09 (95% CI, 0.91–1.31) for anti-S antibodies.

**Table 2.**
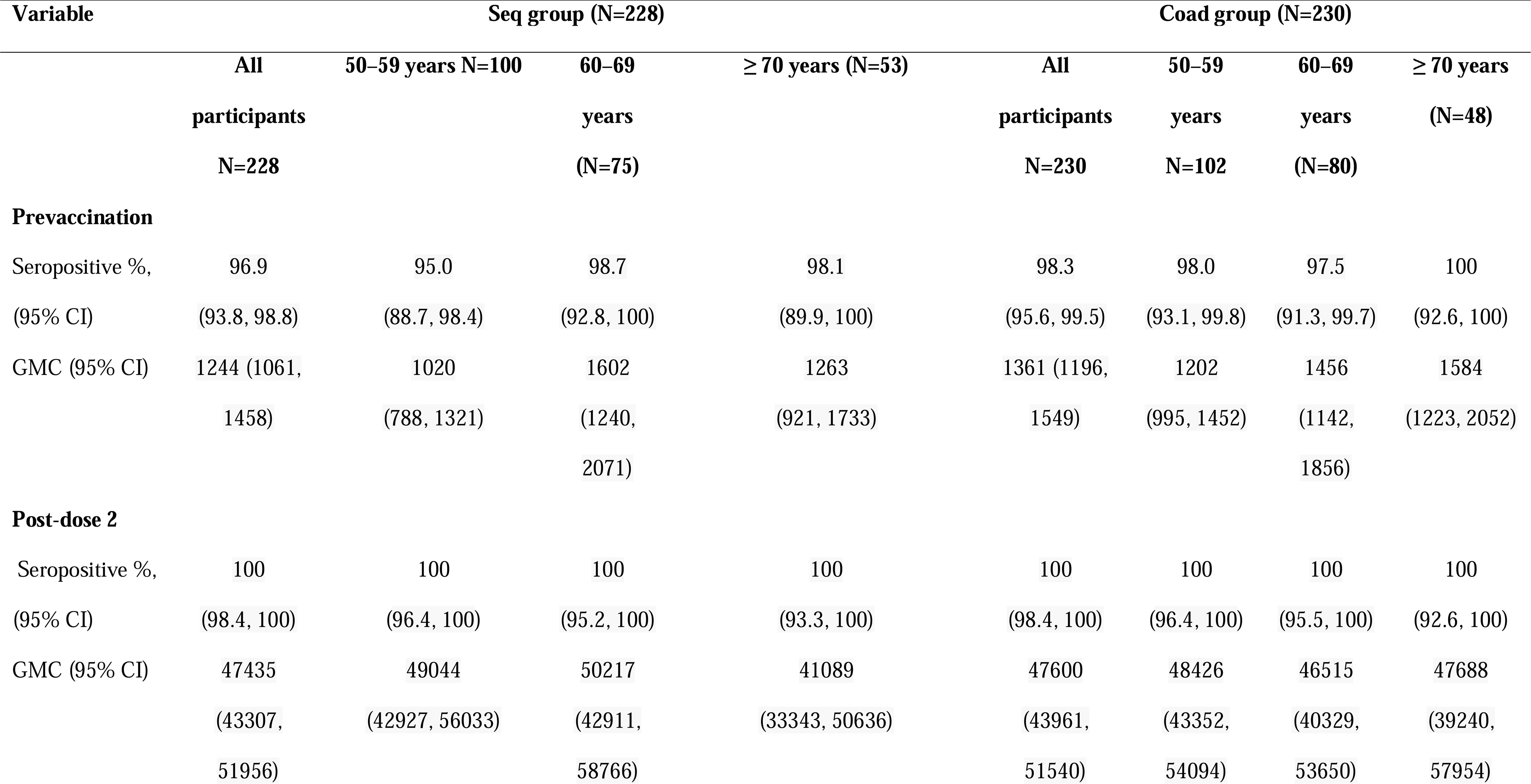
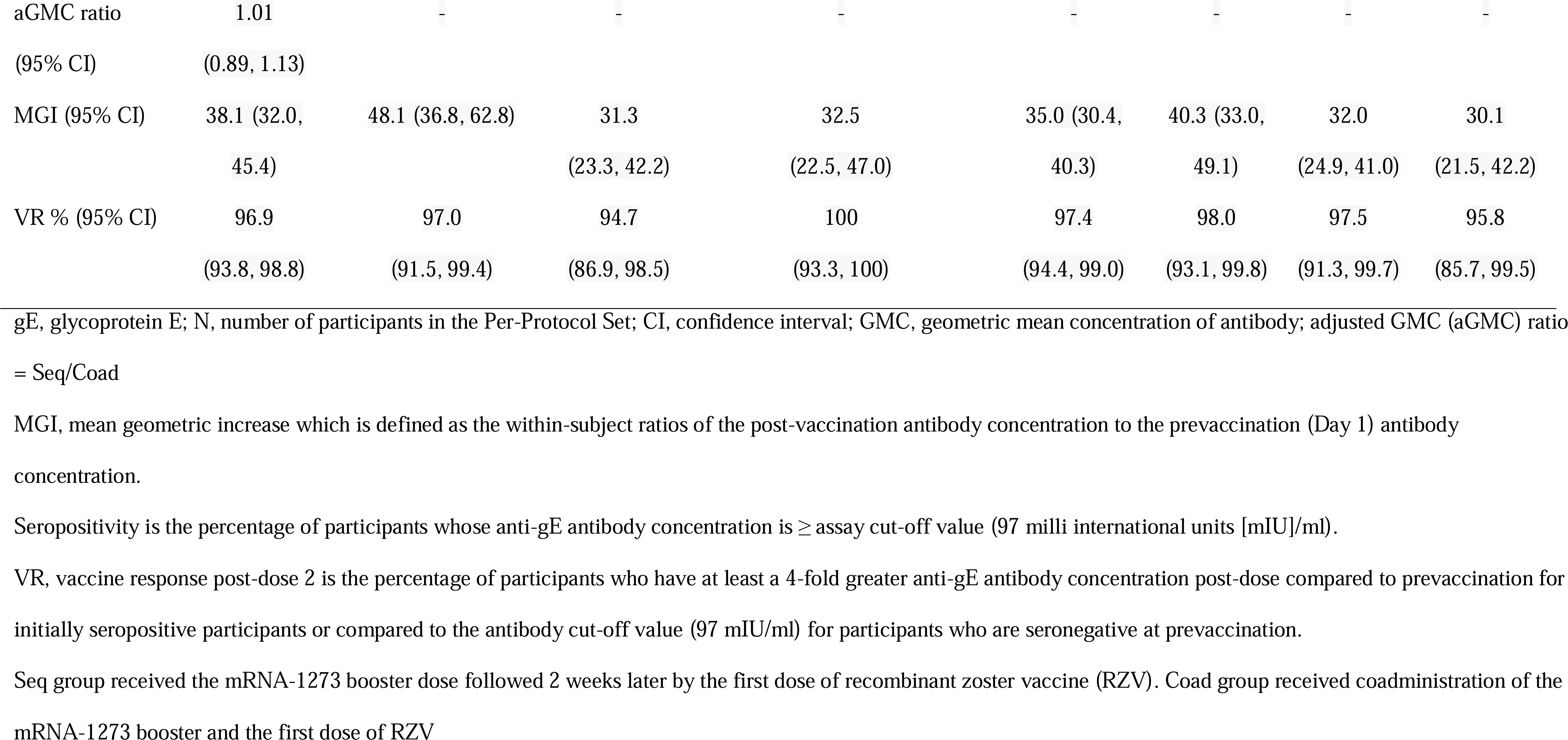
Analysis of anti-gE antibody responses 1 month after the second dose of RZV when the first dose was coadministered with a mRNA-1273 booster dose or administered sequentially 2 weeks later (Per Protocol Set)

**Table 3.**
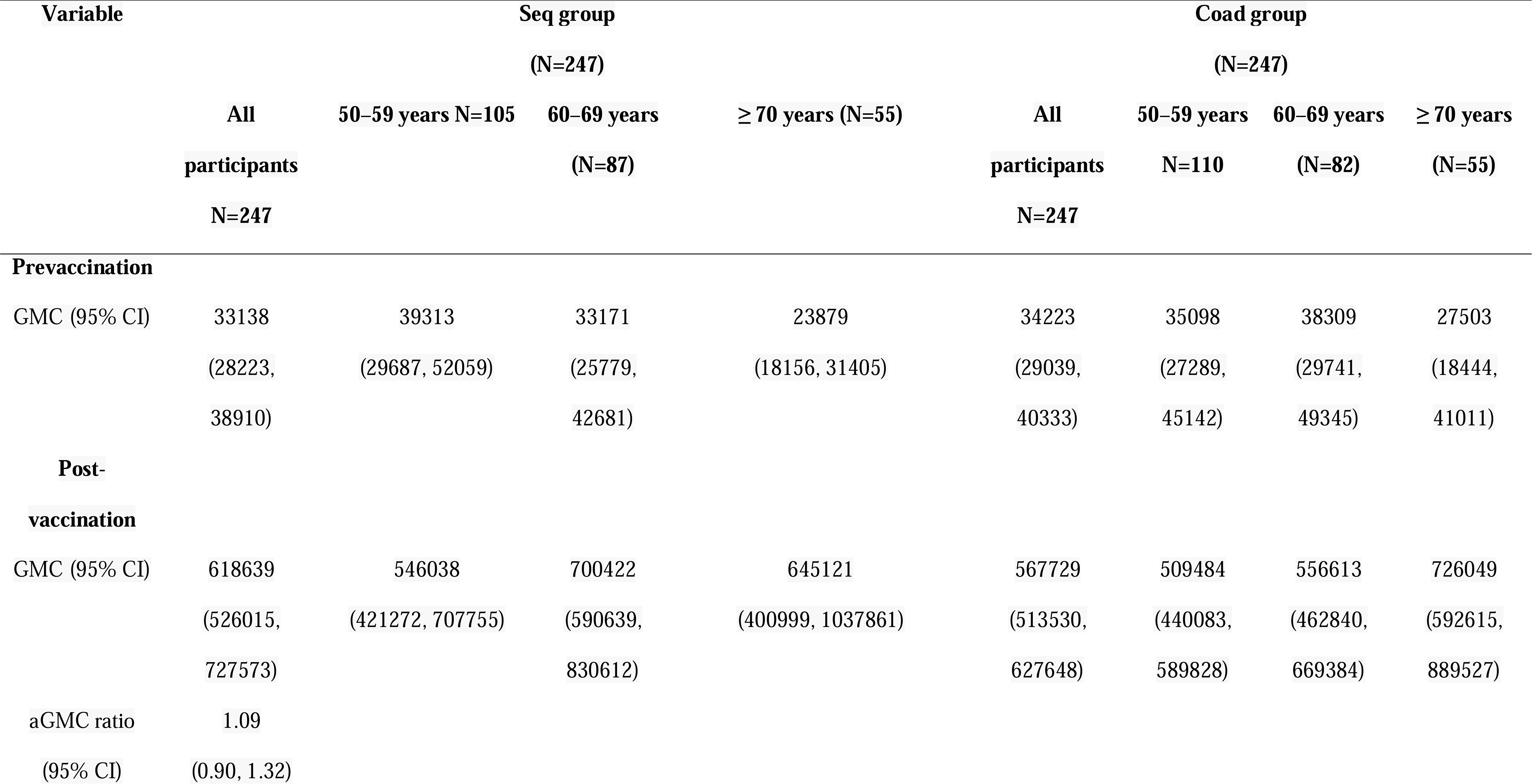
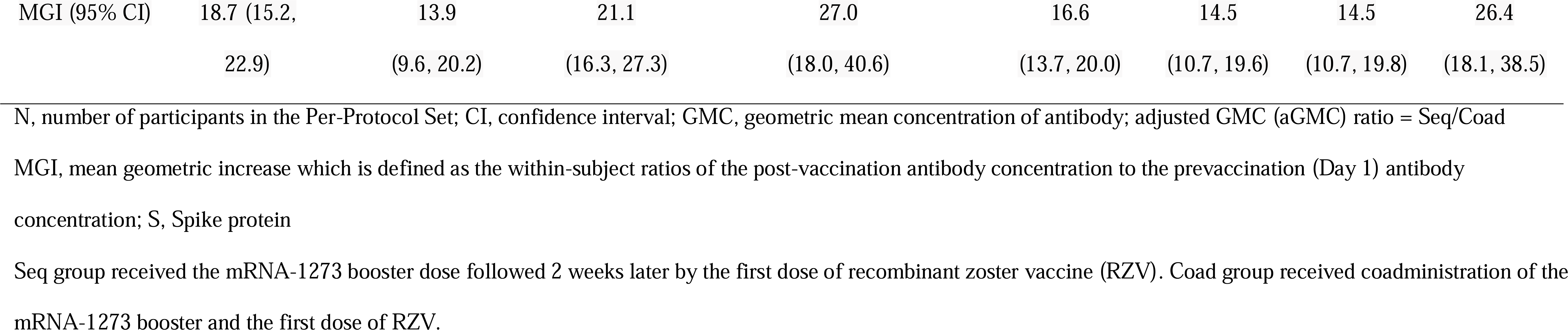
Analysis of anti-S antibody responses 1 month after the mRNA-1273 booster dose when coadministered with the first dose of RZV or when administered sequentially 2 weeks earlier (Per Protocol Set)

Two doses of RZV induced robust antibody responses in all age groups and in both study groups (**Table 2**). One month post-RZV2, the mean geometric increase (MGI) in anti-gE antibodies relative to prevaccination was 38.1 (95% CI, 32.0–45.4) in the Seq group and 35.0 (95% CI, 30.4–40.3) in the Coad group. In both study groups, MGIs tended to be higher in the 50–59-year age group but remained ≥30 in older age groups. Vaccine response rates were ≥94% in both study groups and all age groups.

One month post-mRNA-1273 booster, the MGI in anti-S antibodies was 18.7 (95% CI, 15.2– 22.9) in the Seq group and 16.6 (95% CI, 13.7–20.0) in the Coad group (**Table 3**). MGIs tended to be higher in the ≥70-year age group with lower prevaccination anti-S antibody GMCs than younger adults.

### Safety results

The frequency of any or Grade 3 intensity solicited local AEs post-mRNA-1273 or post- RZV1 were similar in both study groups (**Fig. 2**). The most frequent solicited local AE was injection site pain, both for mRNA-1273 (Seq group 61.8%, Coad group 63.7%) and RZV1 (Seq group 64.1%, Coad group, 70.8%). The median duration of each solicited local AE after mRNA-1273 or RZV1 was from 1 to 2.5 days and similar for both study groups (**Table S1)**. Grade 3 solicited local AEs were uncommon. Grade 3 pain was reported for 1.1% and 1.5% at the mRNA-1273 injection site in the Seq and Coad groups, and at the RZV1 injection site for 1.1% and 2.6% of participants, respectively. The median duration of Grade 3 solicited AEs was 1-3 days and similar in both study groups (**Table S1**).

**Figure 2.**
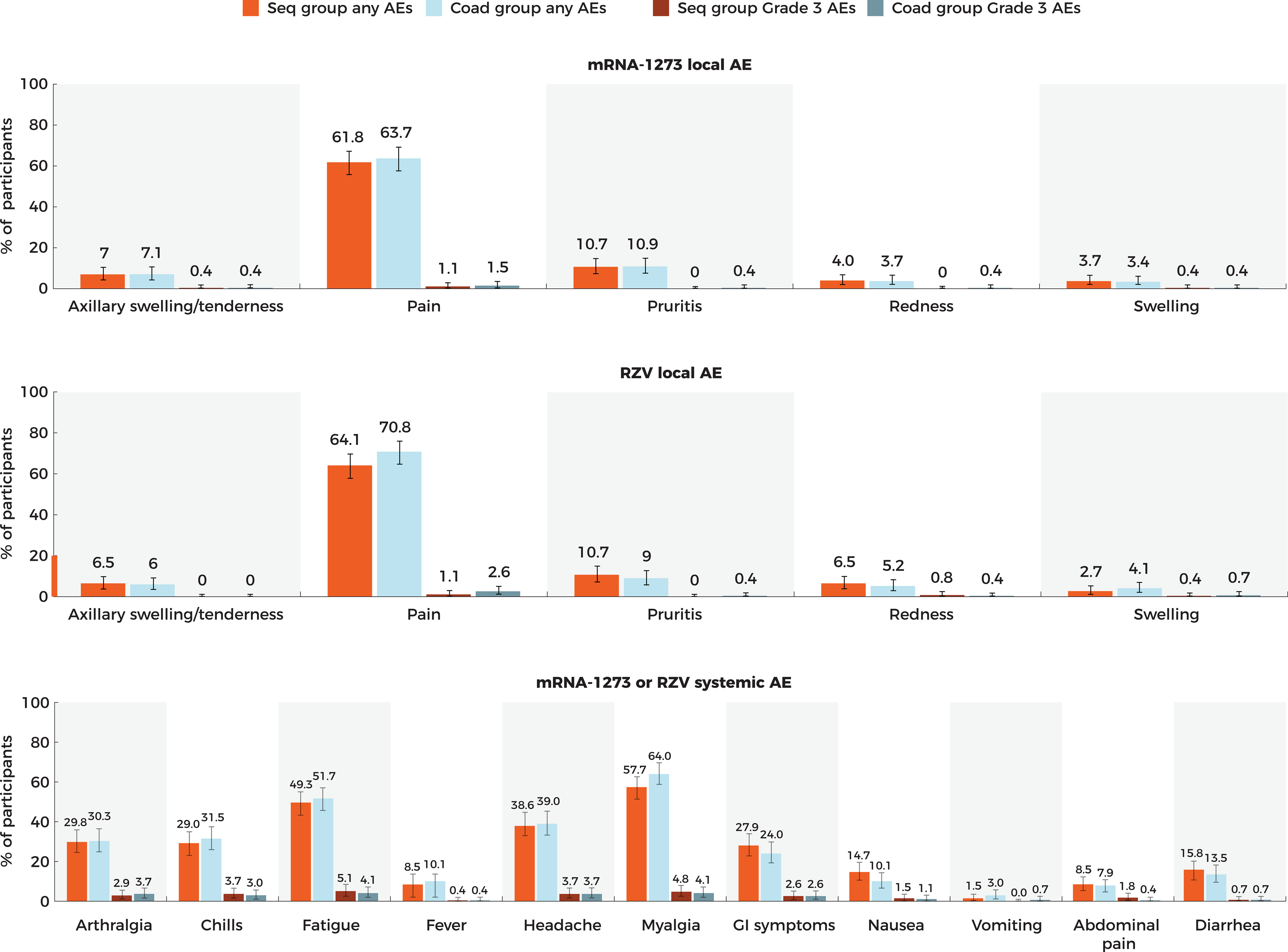
Percentage of solicited local and systemic adverse events reported per participant after the mRNA-1273 and first RZV vaccinations (Exposed Set) AE, adverse event; GI, gastrointestinal; mRNA-1273, Moderna’s mRNA COVID-19 vaccine; RZV, recombinant zoster vaccine Seq group received the mRNA-1273 booster dose followed 2 weeks later by the first dose of RZV. Coad group received coadministration of the mRNA-1273 booster and the first dose of RZV. Definitions of Grade 3 intensity are provided in the Supplement.

The most frequently reported solicited systemic AEs (percentages for Seq and Coad groups, respectively) were myalgia (57.7%, 64.0%), fatigue (49.3%, 51.7%), and headache (38.6%, 39.0%) (**Fig. 2**), and for Grade 3 solicited systemic AEs were fatigue (5.1%, 4.1%) and myalgia (4.8%, 4.1%). The median duration of each solicited systemic AE (any or Grade 3) was from 1 to 1.5 days and similar for both study groups (**Table S1**).

The frequency of solicited AEs of any or Grade 3 intensity reported per participant post- RZV2 was similar in the 2 study groups (**Fig S1**).

There were 41.5% of participants in the Seq group and 46.1% in the Coad group who reported at least 1 unsolicited AE within 30 days of any study vaccination (**Table S2**). Of these, 11/272 (4.0%) in the Seq group and 11/267 (4.1%) in the Coad group had an unsolicited AE assessed by the investigator as related to study vaccination. Related unsolicited AEs reported by at least 1% (n=3) participants in any group were headache (1.1% and 1.5%), fatigue (<1% and 1.1%), diarrhea (0.0% and 1.5%), arthralgia (1.8% and <1%), and myalgia (<1% and 1.1%) in participants in the Seq and Coad groups, respectively. Seven participants (2.6%) in the Seq group and 4 (1.5%) in the Coad group reported Grade 3 unsolicited AEs, of which pulmonary embolism and arthralgia each in 1 participant in the Seq group, and abdominal pain and diarrhea in 1 participant in the Coad group, were assessed by the investigator as related to study vaccination (**Table S3**).

Five participants in the Seq group and 6 in the Coad group reported SAEs. One SAE in the Seq group, pulmonary embolism, in a participant with a history of hyperlipidemia and tobacco use, and with an onset on Day 3 after mRNA-1273 vaccination (prior to receiving RZV1), based on temporal association, was assessed by the investigator and sponsor as related to mRNA-1273 and led to discontinuation from further study vaccinations (**Table S4**). There were no other study vaccine discontinuations due to related AEs.

Three participants in the Seq group and 2 in the Coad group reported AESIs, of which 1 was the pulmonary embolism SAE described above (**Table S4**). An AESI of chronic hepatitis in a participant in the Coad group with a history of rosacea and hypothyroidism, and with onset on Day 36 post-RZV2 was assessed as not related to study vaccines by the investigator, but as the participant was lost to follow-up, based on the limited information available, including liver biopsy results, following a GSK Hepatic Safety panel’s adjudication that the event was possibly drug-induced liver injury with autoimmune features (differential diagnosis included acute hepatitis E), the causality assessment was upgraded by the sponsor to related to both study vaccines.

One participant in each study group reported a pIMD, of which cutaneous vasculitis (no skin biopsy performed) in a participant in the Seq group, with onset on Day 10 after mRNA-1273 and spontaneous resolution prior to administration of RZV1, was assessed as related to mRNA-1273 by the investigator and sponsor based on temporal association (**Table S4**). There were no deaths during the study.

## DISCUSSION

This is the first clinical study to evaluate the safety and immunogenicity of the mRNA-1273 COVID-19 vaccine booster when coadministered with RZV or when administered sequentially. As with sequential administration, coadministration elicited robust anti-gE and anti-S antibody responses, with MGIs exceeding 34-fold and 16-fold, respectively. Notably, in terms of these humoral immune responses, coadministration of the vaccines proved to be noninferior to their sequential administration, confirming an absence of evidence for immune interference upon coadministration. The validity of this result is further strengthened by the same observation of noninferiority from analysis on the Exposed Set. Although for both RZV and mRNA-1273, no validated immunologic mechanistic correlate of protection has been established to date, there is evidence that anti-gE and anti-S antibody responses, as measured in this study, can be considered as reasonable surrogate endpoints likely to predict clinical efficacy for these respective vaccines.^24–27^

The reactogenicity and safety profile of mRNA-1273 and RZV1 coadministration was within their respective Reference Safety Information^18, 28^ and did not reveal any clinically relevant differences versus when the vaccines were administered sequentially.

Potential limitations of the study include the open-label design, which could have influenced safety reporting and causality assessments, although this would be expected to be biased against the Coad group. Coadministration of mRNA-1273 was only assessed post-RZV1; however, since reactogenicity is similar following RZV1 or RZV2 ^16^, one would expect a similar reactogenicity profile if mRNA-1273 was coadministered with RZV2. Finally, the study included a monovalent mRNA vaccine, which has been supplanted by bivalent vaccines; however, these new COVID-19 vaccines containing both emerging and ancestral Spike protein sequences but built on the same mRNA platform are unlikely to induce significantly different immunological mechanisms of action or significantly different safety profiles to those reported herein.^29–31^

In conclusion, the results of this study have shown no safety concerns and a lack of immunologic interference when a booster dose of the COVID-19 mRNA-1273 vaccine was coadministered with the adjuvanted RZV in adults aged ≥50 years. These results provide important data that was previously lacking, to support both the public health agencies whose guidance to health care providers has been that COVID-19 vaccines can be administered without regard to timing of other age-appropriate vaccines, as well as those that are awaiting such data to make evidence-based and unreserved recommendations.^9–11, 14^ It is expected that coadministration of age-appropriate, recommended vaccines at a single clinic visit could be beneficial, especially for older adults, and by increasing coverage rates lead to a decrease in morbidity and mortality due to vaccine preventable diseases.

### Trademark statement

Shingrix is a trademark owned by or licensed to GSK

Spikevax is a trademark of Moderna.

### Contributors

All authors participated in the design or implementation of the study and were involved in the analysis and interpretation of the results and the development of this article. All authors had full access to the data and gave final approval before submission.

### Data sharing

Anonymized individual participant data and study documents can be requested for further research from www.clinicalstudydatarequest.com.

## Acknowledgements

The authors thank the participants, site personnel and investigators who took part in the study. The authors also thank PPD (part of Thermo Fisher Scientific), Moderna, and GSK project team members for their contribution to the study and/or development of the manuscript, especially Julie Vanas (Moderna), Manyee Tsang (GSK), Christophe Dessart (GSK), Raunak Parikh (GSK), Marta Palla (GSK). Business & Decision Life Sciences platform provided editorial assistance and manuscript coordination, on behalf of GSK. Dr Joanne Wolter (independent, on behalf of GSK) provided writing support.

## Plain Language Statement

*What is the context?*

• Booster vaccinations against COVID-19 disease are likely to be necessary for the foreseeable future. Doctors and patients are interested to know whether COVID-19 booster vaccines can be given at the same time as other vaccines in adults.

*What is new?*

• The results of our study showed that an mRNA-based COVID-19 booster vaccine could be coadministered with recombinant shingles vaccine.
• When given together, both vaccines were well tolerated and induced immune responses similar to those observed when they were administered sequentially.

*What is the impact?*

• Administering more than 1 vaccine during a healthcare visit is an efficient way to improve coverage and reduce the number of doctor visits needed to receive all vaccines.
• Coadministration of COVID-19 booster vaccines and the recombinant shingles vaccine could be an attractive option for patients and healthcare professionals.

