## Supplementary material for "No immunological interference or safety concerns when adjuvanted recombinant zoster vaccine is coadministered with a COVID-19 mRNA-1273 booster vaccine in adults aged 50 years and older: A randomized trial"

### Study inclusion criteria

Eligible adults were at least 50 years of age, healthy or medically stable (defined as disease not requiring significant change in therapy or hospitalization for worsening condition for three months before enrolment) and had completed a 2-dose mRNA-1273 primary vaccination series at least 6 months prior to study vaccination according to recommendations in place at the time of study start. Female participants of childbearing potential were required to practice effective contraception for 1 month prior to vaccination through to 2 months after the last dose, and to have a negative pregnancy test prior to each vaccination.

### Study exclusion criteria

Adults were excluded from participation if they had any medical condition that might pose additional risk due to participation in the study, or that might confound post-study safety assessments. Other exclusion criteria were a history of any reaction or hypersensitivity likely to be exacerbated by any study vaccine component; any history of Guillain-Barré syndrome, myocarditis or pericarditis; acute or chronic clinically significant pulmonary, cardiovascular, hepatic or renal functional abnormality or any confirmed or suspected immunosuppressive or immunodeficient condition. Individuals were also excluded if they had hypersensitivity to latex; a history of herpes zoster; had used any investigational or non-registered product up to 30 days before the study vaccination or their planned use during the study period; and if they had received immune-modifying drugs at any time during the study; or immunosuppressants, immunoglobulins and/or any blood products for three months prior to the first dose or planned administration during the study period. Individuals planning to or concurrently participating in another clinical study; or who planned to receive or who had received a vaccine not foreseen by the study protocol from 30 days before dose one until 30 days after the last study vaccination dose were excluded. However, licensed pneumococcal vaccines and non-replicating vaccines were allowed to be administered until eight days prior to dose one and/or dose two of RZV and/or at least 14 days after any dose of RZV. Previous vaccination against herpes zoster (except for live attenuated HZ vaccine) and COVID-19 (except for mRNA-1273 vaccine) were also exclusion criteria.

### Primary and secondary objectives and endpoints relating to coadministration of mRNA-1273 and RZV

| Objectives | Endpoints |
|---|---|
| **Primary** | |
| • To demonstrate the non-inferiority of humoral immunogenicity of 2 doses of RZV when the first dose of RZV is coadministered with the mRNA-1273 booster dose compared to RZV administered alone. | • Anti-glycoprotein E (gE) antibody concentrations expressed as group geometric mean concentration (GMC) ratio at 1 month post-dose 2 of RZV (at Week 14 for RZVSeq, at Week 12 for RZVCoAd). |
| • To demonstrate non-inferiority of humoral immunogenicity of 1 dose of mRNA-1273 booster when the first dose of RZV is coadministered with the mRNA-1273 booster dose compared to mRNA-1273 booster dose administered alone. | • Anti-S Protein antibody concentrations expressed as group GMC ratio at 1 month post mRNA-1273 booster dose (at Week 4 for RZVSeq and RZVCoAd). |
| **Secondary** | |
| • To characterize the anti-gE humoral immunogenicity at prevaccination and at 1 month post-dose 2 of RZV. | • Seropositivity rate with exact 95% CI at prevaccination (at Day 1 for RZVCoAd, at Week 2 for RZVSeq) and at 1 month post-dose 2 of RZV (at Week 14 for RZVSeq, at Week 12 for RZVCoAd).<br>• Anti-gE antibody concentrations expressed as GMC with 95% CI at prevaccination (at Day 1 for RZVCoAd, at Week 2 for RZVSeq) and at 1 month post-dose 2 of RZV (at Week 14 for RZVSeq, at Week 12 for RZVCoAd).<br>• Vaccine response rate (VRR) with exact 95% CIs at 1 month post-dose 2 of RZV (at Week 14 for RZVSeq, at Week 12 for RZVCoAd).<br>• Mean geometric increase (MGI) from prevaccination with 95% CI at 1 month post-dose 2 of RZV (at Week 14 for RZVSeq, at Week 12 for RZVCoAd). |
| • To characterize the humoral immunogenicity of mRNA-1273 booster dose. | • Anti-S Protein antibody concentrations expressed as GMC with 95% CI at prevaccination (Day 1) and 1 month post mRNA-1273 booster dose (at Week 4 for all groups).<br>• MGI from prevaccination with 95% CI at 1 month post mRNA-1273 booster dose (at Week 4 for all groups). |
| • To evaluate the safety and reactogenicity following administration of RZV and mRNA-1273 booster dose, up to 30 days post-last vaccination and during the whole post-vaccination follow-up period. | • Solicited adverse events (AEs): Number and percentage of participants reporting each solicited local AE and each solicited systemic AE within 7 days (Days 1-7) after each dose and overall.<br>• Unsolicited AEs:<br>– Number and percentage of participants reporting unsolicited AEs within 14 days (Days 1-14) after each vaccination visit and overall after any vaccination visit for all groups.<br>– Number and percentage of participants reporting unsolicited AEs within 30 days (Days 1-30) after each vaccination visit and overall after any vaccination visit for all groups.<br>• Serious adverse events (SAEs):<br>– Number and percentage of participants reporting SAEs from first dose up to 30 days post-last dose within each group.<br>– Number and percentage of participants reporting SAEs from first dose to study end.<br>• Potential immune mediated diseases (pIMDs):<br>– Number and percentage of participants reporting of pIMDs from first dose up to 30 days post-last dose within each group. |

| Objectives | Endpoints |
|---|---|
| | – Number and percentage of participants reporting pIMDs from first dose to study end.<br>• Adverse events of special interest (AESIs):<br>– Number and percentage of participants reporting AESIs from first dose up to 30 days post-last dose within each group.<br>– Number and percentage of participants reporting AESIs from first dose to study end.<br>• Suspected HZ episodes: Number and percentage of participants reporting clinically suspected HZ episodes from first dose to study end.<br>• COVID-19 cases: Number and percentage of participants meeting case definitions of COVID-19 from first dose to study end. |

### Study vaccine composition

Each RZV dose contained 50 µg of recombinant gE antigen and the $AS01_B$ adjuvant system (consisting of 50 µg of 3-*O*-desacyl-4′-monophosphoryl lipid A, 50 µg of *Quillaja saponaria* Molina, fraction 21 [licensed by GSK from Antigenics LLC, a wholly owned subsidiary of Agenus Inc., a Delaware, US corporation] and liposome). mRNA-1273 was administered as a 0.25 ml dose containing 50 µg of mRNA embedded in SM-102 lipid nanoparticles. Both vaccines were administered as intramuscular injections, (RZV in the left deltoid and mRNA-1273 in the right deltoid).

### List of adverse events of special interest (AESIs) applicable to mRNA-1273 vaccine

| Medical Concept | Additional Notes |
|---|---|
| **Anosmia, Ageusia** | • New onset COVID associated or idiopathic events without other etiology excluding congenital etiologies or trauma |
| **Subacute thyroiditis** | • Including but not limited to events of: atrophic thyroiditis, autoimmune thyroiditis, immune-mediated thyroiditis, silent thyroiditis, thyrotoxicosis and thyroiditis |
| **Acute pancreatitis** | • Including but not limited to events of: autoimmune pancreatitis, immune-mediated pancreatitis, ischemic pancreatitis, edematous pancreatitis, pancreatitis, acute pancreatitis, hemorrhagic pancreatitis, necrotizing pancreatitis, viral pancreatitis, and subacute pancreatitis<br>• Excluding known etiologic causes of pancreatitis (alcohol, gallstones, trauma, recent invasive procedures) |
| **Appendicitis** | • Include any event of appendicitis |
| **Rhabdomyolysis** | • New onset rhabdomyolysis without known etiology such as excessive exercise or trauma |
| **Acute respiratory distress syndrome (ARDS)** | • Including but not limited to new events of ARDS and respiratory failure. |
| **Coagulation disorders** | • Including but not limited to thromboembolic and bleeding disorders, disseminated intravascular coagulation, pulmonary embolism, deep vein thrombosis |
| **Acute cardiovascular injury** | • Including but not limited to myocarditis, pericarditis, microangiopathy, coronary artery disease, arrhythmia, stress cardiomyopathy, heart failure, or acute myocardial infarction.<br>• Myocarditis or pericarditis |
| **Acute kidney injury** | • Include events with idiopathic or autoimmune etiologies<br>• Exclude events with clear alternate etiology (trauma, infection, tumor, or iatrogenic causes such as medications or radiocontrast etc.)<br>• Include all cases that meet the following criteria:<br>o Increase in serum creatinine by ≥ 0.3 mg/dl (≥26.5 μmol/l) within 48 hours;<br>o OR Increase in serum creatinine to ≥ 1.5 times baseline, known or presumed to have occurred within prior 7 days;<br>o OR Urine volume ≤0.5 ml/ kg/ hour for 6 hours |
| **Acute liver injury** | • Include events with idiopathic or autoimmune etiologies<br>• Exclude events with clear alternate etiology (trauma, infection, tumor, etc.)<br>• Include all cases that meet the following criteria:<br>o > 3-fold elevation above the upper normal limit for alanine aminotransferase or aspartate aminotransferase;<br>o OR > 2-fold elevation above the upper normal limit for total serum bilirubin or gamma-glutamyl transferase or alkaline phosphatase |
| **Dermatologic findings** | • Chilblain-like lesions |

| **Medical Concept** | **Additional Notes** |
|---|---|
| | • Single organ cutaneous vasculitis<br>• Erythema multiforme<br>• Bullous rashes<br>• Severe cutaneous adverse reactions including but not limited to: Stevens-Johnson syndrome, toxic epidermal necrolysis, drug reaction with eosinophilia and systemic symptoms, and fixed drug eruptions |
| **Multisystem inflammatory disorders** | • Multisystem inflammatory syndrome in adults<br>• Multisystem inflammatory syndrome in children<br>• Kawasaki's disease |
| **Thrombocytopenia** | • Platelet counts < 150 $x10^9/l$<br>• Including but not limited to immune thrombocytopenia, platelet production decreased, thrombocytopenia, thrombocytopenic purpura, thrombotic thrombocytopenic purpura, or HELLP (hemolysis, elevated liver enzymes and low platelets) syndrome |
| **Acute aseptic arthritis** | • New onset aseptic arthritis without clear alternate etiology (e.g., gout, osteoarthritis, and trauma) |
| **New onset of or worsening of neurologic disease** | • Including but not limited to:<br>o Guillain-Barré syndrome<br>o Acute disseminated encephalomyelitis<br>o Peripheral facial nerve palsy (Bell's palsy)<br>o Transverse myelitis<br>o Encephalitis/Encephalomyelitis<br>o Aseptic meningitis<br>o Febrile seizures<br>o Generalized seizures/convulsions<br>o Stroke (hemorrhagic and non-hemorrhagic)<br>o Narcolepsy |
| **Anaphylaxis** | • Anaphylaxis |
| **Other syndromes** | • Fibromyalgia<br>• Postural orthostatic tachycardia syndrome<br>• Chronic fatigue syndrome (includes myalgic encephalomyelitis and post viral fatigue syndrome)<br>• Myasthenia gravis |

### Intensity scales for adverse events

| Event | Intensity grade | Parameter |
|---|---|---|
| Pain at administration site | 0 | None |
| | 1 | Mild: Any pain neither interfering with nor preventing normal everyday activities. |
| | 2 | Moderate: Painful when limb is moved and interferes with everyday activities. |
| | 3 | Severe: Significant pain at rest. Prevents normal everyday activities. |
| Redness at administration site | | Greatest surface diameter in mm |
| Swelling at administration site | | Greatest surface diameter in mm |
| Temperature* | | Temperature in °F |
| Axillary (underarm) swelling or tenderness ipsilateral to the side of injection | 0 | None |
| | 1 | Mild: No interference with activity |
| | 2 | Moderate: Repeated use of over-the-counter (non-narcotic) pain reliever >24 hours or some interference with activity |
| | 3 | Severe: Any use of prescription (narcotic) pain reliever or prevents daily activity |
| Pruritis at administration site | 0 | None |
| | 1 | Mild: Itchy sensation that neither interferes with nor prevents normal everyday activities. |
| | 2 | Moderate: Itchy sensation that interferes with normal everyday activities. |
| | 3 | Severe: Itchy sensation that prevents normal everyday activities. |
| Headache | 0 | None |
| | 1 | Mild: Headache that is easily tolerated |
| | 2 | Moderate: Headache that interferes with normal activity |
| | 3 | Severe: Headache that prevents normal activity |
| Fatigue | 0 | None |
| | 1 | Mild: Fatigue that is easily tolerated |
| | 2 | Moderate: Fatigue that interferes with normal activity |
| | 3 | Severe: Fatigue that prevents normal activity |
| Myalgia | 0 | None |
| | 1 | Mild: Myalgia present but does not interfere with activity |
| | 2 | Moderate: Myalgia that interferes with normal activity |
| | 3 | Severe: Myalgia that prevents normal activity |
| Arthralgia | 0 | None |
| | 1 | Mild: Arthralgia present but does not interfere with activity |
| | 2 | Moderate: Arthralgia that interferes with normal activity |
| | 3 | Severe: Arthralgia that prevents normal activity |
| Gastrointestinal symptoms | 0 | None |
| | 1 | Mild: Gastrointestinal symptoms that are easily tolerated |
| | 2 | Moderate: Gastrointestinal symptoms that interfere with normal activity |
| | 3 | Severe: Gastrointestinal symptoms that prevent normal activity |
| Shivering/chills | 0 | None |
| | 1 | Mild: Shivering that is easily tolerated |
| | 2 | Moderate: Shivering that interferes with normal activity |
| | 3 | Severe: Shivering that prevents normal activity |

*Fever is defined as temperature ≥ 38.0°C (100.4°F) by any route. The preferred location for measuring temperature will be the oral route.

The maximum intensity of injection administration site redness/swelling and fever was graded as follows:

| | Redness/swelling (diameter) | Fever |
|---|---|---|
| 0: | ≤20 mm | <38.0°C (100.4°F) |
| 1: | >20 - ≤50 mm | ≥38.0°C (100.4°F) - ≤38.5°C (101.3°F) |
| 2: | >50 - ≤100 mm | >38.5°C (101.3°F) - ≤39.0°C (102.2°F) |
| 3: | >100 mm | >39.0°C (102.2°F) |

The investigator assessed the maximum intensity that occurred over the duration of the event for all unsolicited adverse events recorded during the study. The intensity was assigned to one of the following categories:

| | | |
|---|---|---|
| 1 (mild) | = | An adverse event (AE) which is easily tolerated by the participant, causing minimal discomfort and not interfering with everyday activities. |
| 2 (moderate) | = | An AE which is sufficiently discomforting to interfere with normal everyday activities. |
| 3 (severe) | = | An AE which prevents normal, everyday activities. Such an AE would, for example, prevent attendance at work/school and would cause the participant to seek medical advice. |

### Assessment of causality

Causality was assessed by the investigator using the following question: *Is there a reasonable possibility that the unsolicited AE may have been caused by the study intervention?*

YES: There is a reasonable possibility that the study intervention contributed to the AE.

NO: There is no reasonable possibility that the AE is causally related to the administration of the study intervention. There are other, more likely causes and administration of the study intervention is not suspected to have contributed to the AE.

### Study holding rules

Randomization was to be temporarily put on hold and an *ad hoc* SRT review performed if any of the holding rule criteria were met at any time during the study. Randomization could only be resumed following SRT approval.

| **Holding Rule** | **Event** | **Number or percentage of participants** |
|---|---|---|
| | | **Coad group** |
| 1a | Death or any life-threatening SAE that can be reasonably attributed to the vaccination | 1 participant |
| 1b | Any non-life-threatening SAE that can be reasonably attributed to the vaccination | 3/28 participants (for initial SRT review);<br>For the rest of the study: 10% |
| 1c | Any withdrawal from the study (by investigator or participant request) following a Grade 3 AE that can be reasonably attributed to the vaccination | 3/28 participants (for initial SRT review);<br>For the rest of the study: 10% |
| 1d | Fever >40°C (104°F), OR<br>Any solicited local event or solicited systemic AE leading to hospitalization, each within the 7-day (Days 1–7) post-vaccination period | 2 participants |
| 2a | Any Grade 3 solicited systemic AE (lasting 48 hours or more) within the 7-day (Day 1–7) post-vaccination period | 6/28 participants (for initial SRT review);<br>For the rest of the study: 20% |
| 2b | Any Grade 3 unsolicited AE, that can be reasonably attributed to the vaccination, within the 7-day (Day 1–7) post-vaccination period | 3/28 participants (for initial SRT review);<br>For the rest of the study: 10% |

AE, adverse event; SAE, serious adverse event; SRT, safety review team

**Figure S1 Percentage of participants reporting solicited local and systemic adverse events after the second RZV vaccination**

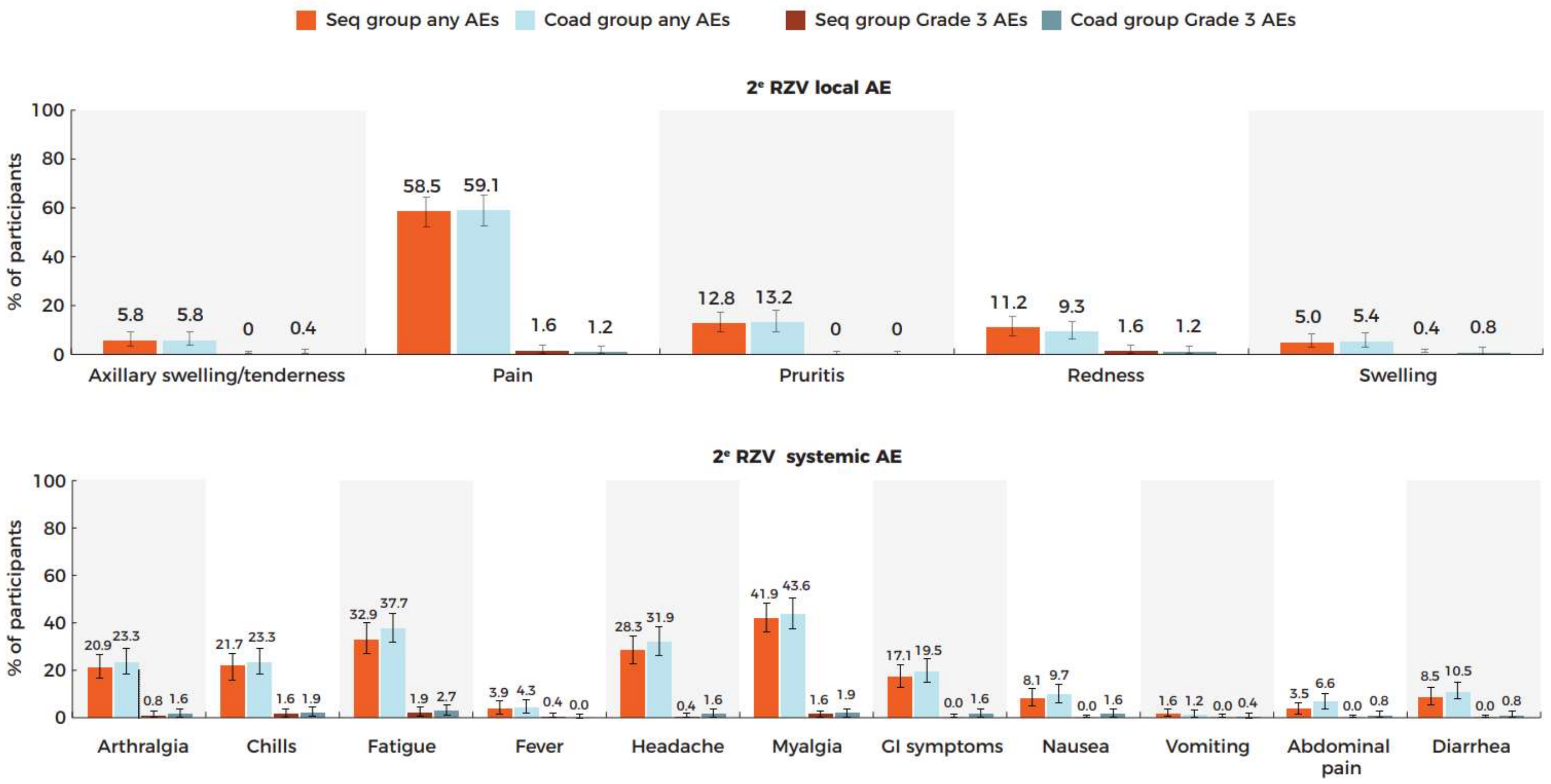

AE, adverse event; GI, gastrointestinal; mRNA-1273, Moderna's mRNA COVID-19 vaccine; RZV, recombinant zoster vaccine
Seq group received the mRNA-1273 booster dose followed 2 weeks later by the first dose of RZV. Coad group received coadministration of the mRNA-1273 booster and the first dose of RZV. Both groups received the second dose of RZV 2 months later
Definitions of Grade 3 intensity are provided above.

### Table S1 Median duration (and range) in days of solicited adverse events

| | Seq group (N=272) | | Coad group (N=267) | |
|---|---|---|---|---|
| | **mRNA-1273 injection site** | **RZV1 injection site** | **mRNA-1273 injection site** | **RZV1 injection site** |
| **Solicited local reactions** | | | | |
| Any | | | | |
| Pain | 2.0 (1-7) | 2.0 (1-10) | 2.0 (1-8) | 2.0 (1-10) |
| Axillary swelling/tenderness | 2.0 (1-6) | 1.0 (1-3) | 2.0 (1-5) | 1.0 (1-5) |
| Pruritis | 1.0 (1-5) | 1.5 (1-5) | 1.0 (1-9) | 1.0 (1-10) |
| Redness | 2.0 (1-5) | 2.0 (1-5) | 2.5 (1-6) | 1.0 (1-3) |
| Swelling | 2.0 (1-5) | 2.0 (1-4) | 1.0 (1-5) | 1.0 (1-3) |
| Grade 3 | | | | |
| Pain | 1.0 (1-1) | 1.0 (1-1) | 1.0 (1-3) | 1.0 (1-3) |
| Axillary swelling/tenderness | 1.0 (1-1) | 0 | 2.0 (2-2) | 0 |
| Pruritis | 0 | 0 | 1.0 (1-1) | 1.0 (1-1) |
| Redness | 0 | 3.0 (1-5) | 1.0 (1-1) | 1.0 (1-1) |
| Swelling | 1.0 (1-1) | 2.0 (2-2) | 1.0 (1-1) | 1.0 (1-1) |
| **Solicited systemic reactions** | **mRNA-1273** | **RZV1** | **mRNA-1273 and RZV1** | |
| Any | | | | |
| Abdominal pain | 1.0 (1-23) | 1.0 (1-1) | 1.0 (1-12) | |
| Arthralgia | 1.0 (1-6) | 1.0 (1-7) | 1.0 (1-55) | |
| Chills | 1.0 (1-3) | 1.0 (1-4) | 1.0 (1-3) | |
| Diarrhea | 1.0 (1-3) | 1.5 (1-8) | 1.0 (1-11) | |
| Fatigue | 1.0 (1.5) | 1.0 (1-7) | 1.0 (1-13) | |
| Fever | 1.0 (1-2) | 1.0 (1-1) | 1.0 (1-2) | |
| Gastrointestinal symptoms | 1.0 (1-23) | 1.0 (1-8) | 1.0 (1-12) | |
| Headache | 1.0 (1-22) | 1.0 (1-4) | 1.0 (1-11) | |
| Myalgia | 1.0 (1-6) | 1.0 (1-6) | 1.0 (1-55) | |
| Nausea | 1.0 (1-3) | 1.0 (1-2) | 1.0 (1-3) | |
| Vomiting | 1.0 (1-1) | 0 | 1.0 (1-1) | |
| Grade 3 | | | | |
| Abdominal pain | 1.5 (1-21) | 1.0 (1-1) | 1.0 (1-1) | |
| Arthralgia | 1.0 (1-2) | 1.0 (1-1) | 1.0 (1-3) | |
| Chills | 1.0 (1-2) | 1.0 (1-1) | 1.0 (1-2) | |
| Diarrhea | 1.0 (1-1) | 0 | 1.0 (1-1) | |
| Fatigue | 1.0 (1-2) | 1.0 (1-1) | 1.0 (1-3) | |
| Fever | 0 | 1.0 (1-1) | 1.0 (1-1) | |
| Gastrointestinal symptoms | 1.5 (1-21) | 1.0 (1-1) | 1.0 (1-1) | |
| Headache | 1.0 (1-2) | 1.0 (1-1) | 1.0 (1-2) | |
| Myalgia | 1.0 (1-2) | 1.0 (1-1) | 1.0 (1-3) | |
| Nausea | 1.0 (1-1) | 1.0 (1-1) | 1.0 (1-1) | |
| Vomiting | 0 | 0 | 1.0 (1-1) | |

N, number of participants in respective study group who have any diary data entered; RZV1, first dose of recombinant zoster vaccine

Duration is the number of days on which participant experienced the adverse event.

Seq group received the mRNA-1273 booster dose followed 2 weeks later by the first dose of recombinant zoster vaccine (RZV). Coad group received coadministration of the mRNA-1273 booster and the first dose of RZV

### Table S2 Unsolicited adverse events within 30 days after any study vaccination reported by at least 1% of participants in any group

| | Seq group (N=272) | | Coad group (N=267) | |
|---|---|---|---|---|
| **Preferred Term** | **n (%)** | **95% CI** | **n (%)** | **95% CI** |
| **At least 1 unsolicited AE within 30 days** | 113 (41.5%) | 35.6, 47.6 | 123 (46.1%) | 40.0, 52.2 |
| Headache | 49 (18.0%) | 13.6, 23.1 | 62 (23.2%) | 18.3, 28.8 |
| Fatigue | 32 (11.8%) | 8.2, 16.2 | 48 (18.0%) | 13.6, 23.1 |
| Chills | 15 (5.5%) | 3.1, 8.9 | 18 (6.7%) | 4.0, 10.4 |
| Diarrhoea | 27 (9.9%) | 6.6, 14.1 | 40 (15.0%) | 10.9, 19.8 |
| Abdominal pain | 13 (4.8%) | 2.6, 8.0 | 21 (7.9%) | 4.9, 11.8 |
| Nausea | 16 (5.9%) | 3.4, 9.4 | 15 (5.6%) | 3.2, 9.1 |
| Vomiting | 4 (1.5%) | 0.4, 3.7 | 3 (1.1%) | 0.2, 3.2 |
| Arthralgia | 31 (11.4%) | 7.9, 15.8 | 28 (10.5%) | 7.1, 14.8 |
| Myalgia | 26 (9.6%) | 6.3, 13.7 | 27 (10.1%) | 6.8, 14.4 |
| COVID-19 | 3 (1.1%) | 0.2, 3.2 | 5 (1.9%) | 0.6, 4.3 |
| Urinary tract infection | 2 (<1%) | 0.1, 2.6 | 5 (1.9%) | 0.6, 4.3 |
| Upper respiratory tract infection | 5 (1.8%) | 0.6, 4.2 | 1 (<1%) | 0.0, 2.1 |
| Procedural pain | 0 (0.0%) | 0.0, 1.3 | 3 (1.1%) | 0.2, 3.2 |
| **Any related unsolicited AE within 30 days** | | | | |
| Fatigue | 1 (<1%) | 0.0, 2.0 | 3 (1.1%) | 0.2, 3.2 |
| Headache | 3 (1.1%) | 0.2, 3.2 | 4 (1.5%) | 0.4, 3.8 |
| Diarrhoea | 0 (0.0%) | 1.0, 1.3 | 4 (1.5%) | 0.4, 3.8 |
| Arthralgia | 5 (1.8%) | 0.6, 4.2 | 2 (<1%) | 0.1, 2.7 |
| Myalgia | 1 (<1%) | 0.0, 2.0 | 3 (1.1%) | 0.2, 3.2 |

AE, adverse event; CI, confidence interval; COVID-19, coronavirus disease 2019; N, number of participants in each group; n, number of participants with the indicated adverse event.

Seq group received the mRNA-1273 booster dose followed 2 weeks later by the first dose of recombinant zoster vaccine (RZV). Coad group received coadministration of the mRNA-1273 booster and the first dose of RZV

### Table S3 Unsolicited adverse events of Grade 3 intensity within 30 days after any study vaccination

| | Seq group (N=272) | | Coad group (N=267) | |
|---|---|---|---|---|
| **Preferred Term** | **n (%)** | **95% CI** | **n (%)** | **95% CI** |
| **Any Grade 3 unsolicited AE within 30 days** | 7 (2.6%) | 1.0, 5.2 | 4 (1.5%) | 0.4, 3.8 |
| COVID-19 | 1 (<1%) | 0.0, 2.0 | 0 (0.0%) | 0.0, 1.4 |
| Urosepsis | 1 (<1%) | 0.0, 2.0 | 0 (0.0%) | 0.0, 1.4 |
| Arthralgia | 2 (<1%) | 0.1, 2.6 | 0 (0.0%) | 0.0, 1.4 |
| Basal cell carcinoma | 1 (<1%) | 0.0, 2.0 | 0 (0.0%) | 0.0, 1.4 |
| Intraductal proliferative breast lesion | 0 (0.0%) | 0.0, 1.3 | 1 (<1%) | 0.0, 2.1 |
| Abdominal pain | 0 (0.0%) | 0.0, 1.3 | 1 (<1%) | 0.0, 2.1 |
| Diarrhoea | 0 (0.0%) | 0.0, 1.3 | 1 (<1%) | 0.0, 2.1 |
| Ankle fracture | 1 (<1%) | 0.0, 2.0 | 0 (0.0%) | 0.0, 1.4 |
| Anxiety | 0 (0.0%) | 0.0, 1.3 | 1 (<1%) | 0.0, 2.1 |
| Pulmonary embolism | 1 (<1%) | 0.0, 2.0 | 0 (0.0%) | 0.0, 1.4 |
| Peripheral venous disease | 0 (0.0%) | 0.0, 1.3 | 1 (<1%) | 0.0, 2.1 |
| **Any related Grade 3 unsolicited AE within 30 days** | 2 (<1%) | 0.1, 2.6 | 1 (<1%) | 0.0, 2.1 |
| Arthralgia | 1 (<1%) | 0.0, 2.0 | 0 (0.0%) | 0.0, 1.4 |
| Abdominal pain | 0 (0.0%) | 0.0, 1.3 | 1 (<1%) | 0.0, 2.1 |
| Diarrhoea | 0 (0.0%) | 0.0, 1.3 | 1 (<1%) | 0.0, 2.1 |
| Pulmonary embolism | 1 (<1%) | 0.0, 2.0 | 0 (0.0%) | 0.0, 1.4 |

AE, adverse event; CI, confidence interval; COVID-19, coronavirus disease 2019; N, number of participants in each group; n, number of participants with the indicated adverse event.
Seq group received the mRNA-1273 booster dose followed 2 weeks later by the first dose of recombinant zoster vaccine (RZV). Coad group received coadministration of the mRNA-1273 booster and the first dose of RZV.

### Table S4 AESIs, pIMDs and SAEs reported from the first study vaccination until study end

| | Seq group (N=272) | | Coad group (N=267) | |
|---|---|---|---|---|
| **Preferred Term** | **n (%)** | **95% CI** | **n (%)** | **95% CI** |
| **At least 1 AESI until study end** | 3 (1.1%) | | 2 (<1%) | |
| Pancreatitis acute | 0 (0.0%) | | 1 (<1%) | |
| Chronic hepatitis | 0 (0.0%) | | 1 (<1%) | |
| Seizure | 1 (<1%) | | 0 (0.0%) | |
| Pulmonary embolism | 1 (<1%) | | 0 (0.0%) | |
| Cutaneous vasculitis | 1 (<1%) | | 0 (0.0%) | |
| **At least 1 pIMD until study end** | 1 (<1%) | | 1 (<1%) | |
| Gout | 0 (0.0%) | | 1 (<1%) | |
| Cutaneous vasculitis | 1 (<1%) | | 0 (0.0%) | |
| **At least 1 SAE until study end** | 5 (1.8) | 0.6, 4.2 | 6 (2.2) | 0.8,4.8 |
| Pulmonary embolism | 1 | | | |
| Ankle fracture | 1 | | | |
| Seizure | 1 | | | |
| Urosepsis | 1 | | 1 | |
| Hypoglycemia | 1 | | | |
| Nephrolithiasis | | | 1 | |
| Hydronephrosis | | | 1 | |
| Pancreatitis acute | | | 1 | |
| Intraductal proliferative breast lesion | | | 1 | |
| Osteoarthritis | | | 1 | |
| Cellulitis | | | 1 | |
| Endometrial cancer | | | 1 | |

AESI, adverse event of special interest; CI, confidence interval; N, number of participants in each group; n, number of participants with the indicated adverse event; pIMD, potential immune-mediated disease; SAE, serious adverse event.
Seq group received the mRNA-1273 booster dose followed 2 weeks later by the first dose of recombinant zoster vaccine (RZV). Coad group received coadministration of the mRNA-1273 booster and the first dose of RZV.
